# Drivers of youth engagement in mental health in Morocco: findings from a nationwide cross-sectional survey

**DOI:** 10.1101/2025.04.12.25325717

**Authors:** Fatima Ezzahraa Wafqui, Oumnia Bouaddi, Imad Elbadisy, Mohamed Khalis

## Abstract

**Background:** Youth engagement in mental health has been shown to inform effective interventions aimed at improving youth mental health outcomes. However, evidence on the state of youth engagement in mental health remains limited in low- and middle-income countries (LMICs). This study aims to identify the drivers of youth engagement in mental health in Morocco, as well as the support needs and resources required to promote it.

**Methods:** We conducted a nationwide cross-sectional study in Morocco, including young Moroccans aged 18–24 years. Using an online self-administered questionnaire, we assessed participants’ levels of engagement in mental health activities, attitudes toward mental health, awareness about mental health, support needs, and perceived importance of well-being drivers across five domains, informed by the WHO adolescent well-being framework. Descriptive statistics, clustering analysis, and logistic regression analyses were used to identify predictor factors of youth engagement in mental health.

**Results:** A total of 1,183 participants were included. The engaged cluster group reported higher awareness about mental health and more positive attitudes toward mental health. Predictors of engagement in mental health included higher education (OR=2.23, 95% CI: [46-3.43]) unemployment (OR=1.65, 95% CI: [1.04-2.64]), and higher scores on “Connectedness and positive values and contribution to society” (OR=1.07, 95% CI: [1.05-1.10]) as well as positive attitudes toward mental health (OR=1.04, 95% CI: [1.03-1.05]). Conversely, those who prioritized ‘safety and a supportive environment’ were less likely to be engaged (OR=0.94, 95% CI: [0.91-0.98]). The most frequently cited needs to support engagement were access to mental health professionals (63.0%) and mental health education (42.4%).

**Conclusions:** These results provide insights into the factors influencing youth engagement in mental health in Morocco. Fostering “connectedness, positive values, and contribution to society” and “positive attitudes towards mental health”, as well as improving access to mental health professionals, information and education, is essential to promote youth engagement in mental health programs and policy-making.

## 1. Introduction

Mental health issues are considered the Non-Communicable Diseases (NCDs) of the young given their early onset and their extension to adulthood (1). Globally, estimates show that 50% of young people will have experienced at least one period of poor mental health by the age of 25 with 75% of mental disorders having an onset before that age (1). Given the economic and deleterious consequences of lack to access to mental health services (2), the World Health Organization (WHO) has calls for the promotion of child and adolescent mental health (CAMH) (3) as best buy for the Future Mental Capital of Individuals, Communities and Economies (4). Despite these global challenges to advance mental health outcomes remain and all countries are considered “developing” with regards to prioritizing mental health (5).

In Morocco, young people aged up to 17 years old represented 31.5% of the total population in 2022, while young adults aged 15 to 29 years old represented almost half of the working age population (6,7). In 2019, a report by the High Commission of Planning (HCP) showed that Moroccan youth often deal with significant stressors at home, ranging from domestic violence, household instability and authoritative parenting style (8). In addition, the absence of adequate safe spaces exacerbates the mental health struggles of the young, contributing to more psychological distress, life long struggle with mental health issues (9). According to the Global School-Based Health Survey (GSHS) in 2016, 16% of Moroccan students aged surveyed were found to have seriously considered committing suicide (10). This phenomenon was more common among girls compared to boys (17.9% vs. 14%). Additionally, young people attending schools and universities perceive these environments as spaces where they encounter adverse experiences such as an academic culture solely focused on performance, bullying, violence, harassment and somewhat easy access to harmful substances (8). In 2017, more than 60% of youth aged 13 to 17 years old who smoked or drank alcohol, have taken up this behaviour before the age of 14 (11). The National Strategic Plan for Mental Health Promotion in Children, Adolescents and Young People, and the National Strategy for Youth Health in Morocco recognizes mental health promotion as indispensable for the reaching SDG3 and efforts to reinforce prevention and treatment of substance use and implementation of a universal health coverage are currently underway including promotion of access to mental health services, creation of university health services and units, and the setting up of some counselling services within educational institutions (12). Despite these efforts, a digital citizen consultation of more than 27,000 in 2022 found that more than 75% were little or not informed about public youth programs; and early considered these programs for young people to be unsuccessful (13).

An extensive body of literature shows that engaging youth in mental health efforts and initiatives is a powerful means to directly and indirectly contribute to their mental health and well-being (14). However, the extent to which youth are included in mental health programming remains unknown, and very little research has been conducted in LMIC contexts to explore this issue. A consultation of stakeholder perspectives in multiple LMICs, including Morocco, revealed that many young people expressed frustration with the lack of power-sharing by adults and feelings of being accessories which is a barrier to meaningful youth participation in mental health policymaking (15). In addition, marginalized groups were found to be less likely to be engaged in mental health programs. Building on this knowledge, and given that the factors driving youth engagement in mental health have not been previously documented or explored in the Moroccan context, this paper aims to identify the level of youth engagement in mental health in Morocco and explore the predictive factors influencing such engagement

## 2. Methods

### 2.1. Study design and setting

We conducted a nationwide cross-sectional online survey among Moroccan young people aged 18 to 24 years across the 12 regions of Morocco. This study took place from November 2023 and February 2024.

### 2.2. Study population and sampling

We used stratified random sampling to allocate the sample size across regions and sub-regions, considering sociodemographic characteristics such as place of residence, sex, and level of education. First, we proportionally allocated the number of participants to each region and then applied simple random sampling within each region. We included young people aged 18 to 24 years from diverse sociodemographic backgrounds residing in any of Morocco’s 12 regions.

### 2.3. Data collection

We trained twelve young people, one from each of the 12 regions, to support participant recruitment and data collection. These individuals were selected for their active roles in civil society and community organizations, which ensured strong connections to the target population. The survey was distributed using WhatsApp. Data collection was conducted using Microsoft Forms via a self-administered online survey available in both Arabic and French. The survey was self-administered for computer-literate participants and administered by the trained youth for the remaining participants. The questionnaire included five sections. The first section collected participant sociodemographic characteristics and general information (age, gender, residency, marital status, education, occupation). The second section, was informed by the WHO Adolescent Wellbeing Framework, and included five domains underlying youth and adolescent well-being (16) : (i) good health and optimum nutrition, (ii) connectedness, positive values, and contribution to society, (iii) safety and a supportive environment, (iv) learning, competence, education, skills, and employability, and (v) agency and resilience. The third section assessed awareness about mental health and the fourth section, collected assessed attitudes towards mental health using items available in the literature. The final section, collected information about youth engagement (level of engagement, and support needed to be more engaged in initiatives or activities related to mental health). The English version of the questionnaire is available in supplementary material.

For each of the five well-being domains, the following scores were calculated by using a 5-point Likert scale, ranging from 0 (“Not important at all”) to 4 (“Extremely important”): Score of good health and optimum nutrition (nine items, possible range = 0 – 36 points), score of connectedness, positive values, and contribution to society (nine items, possible range = 0 – 36 points), score of safety and a supportive environment (seven items, possible range = 0 – 28 points), score of learning, competence, education, skills, and employability (eight items, possible range = 0 – 32 points), and score of agency and resilience (six items, possible range = 0 – 24 points).

We summarized awareness about mental health by calculating a score based on the eight items, this score ranged from 0 (low level of awareness) to 32 points (high level of awareness). We summarized attitudes towards mental health by calculating a score based on the thirteen items, this score ranged from 0 (negative attitude) to 52 points (positive attitude).

We assessed the level of youth engagement based on six following items: frequency of engagement in activities related to mental health and well-being, involvement in the design of programs and activities related to mental health, involvement in the implementation of mental health programs, involvement in keeping track of and evaluating mental health programs, actively involved in raising awareness about mental health, and participating in groups that make decisions about mental health. For each item, participants were asked to rate their engagement on a 5-point Likert scale, where “0” is never” and “4” is always. We assigned engagement score to participants by summing the individual scores for each item, consequently, the total score of engagement ranges between 0 (low engagement) and 24 (high engagement).

The survey was piloted among 20 participants who met the inclusion criteria with feedback used to edit and modify the survey prior to its use for data collection.

### 2.4. Statistical analysis

Data was entered into Excel sheets and cleaned, where it was checked for validity and completeness. Descriptive analyses were conducted using frequencies, percentages for categorical variables, and means (± standard deviation) for continuous variables. The level of youth engagement (dependent variable) was used as a binary variable (engaged / not engaged) using median value of the score as cut-off point.

To identify distinct youth profiles based on key indicators of mental health and well-being, we applied a clustering approach suitable for mixed-type data, comprising both categorical and numerical variables. All character-type variables were initially converted into factors to ensure appropriate handling during dissimilarity computation. A pairwise dissimilarity matrix was then computed using Gower’s distance (17). Clustering was subsequently performed using the k-means algorithm, applied to the Gower distance matrix (18). The number of clusters was initially set to k = 3, based on theoretical considerations regarding youth heterogeneity, and further evaluated using the elbow method, which assesses the within-cluster sum of squares to identify the point at which additional clusters provide minimal gain in explained variance (19). Each individual was assigned to one of the three clusters, and the dataset was augmented with cluster membership labels. Subsequent descriptive analyses were conducted to examine the distribution of sociodemographic and psychosocial variables across clusters. This clustering process aimed to uncover latent groupings among youth, revealing diverse patterns of engagement and mental health-related attributes.

The crude and adjusted odds ratios (OR), and their 95% confidence intervals, of factors associated with engagement, were estimated using logistic regression models. Multivariate adjusted odds ratios were calculated to adjust for age, gender, residency, marital status, education, occupation, score of good health and optimum nutrition, score of connectedness, positive values, and contribution to society, score of safety and a supportive environment, score of learning, competence, education, skills, and employability, score of agency and resilience, score of awareness about mental health, and score of attitudes towards mental health. All statistical analyses were conducted using R software (version 4.4.0).

### 2.6. Ethical considerations

Approval for the study protocol was granted by the Ethics Committee of the Faculty of Medicine and Pharmacy of Tangier under number (AC470C/2023). Participation was voluntary and electronic informed consent was obtained from all participants before data collection began. Confidentiality was maintained and no identifiable information was collected from participants.

## Results

### 3.1. Socio-demographic characteristics of Study Population

A total of 1,183 participants aged 18-24 years old were included in the final analysis (mean age 20.5 ±2.0) (Table 1). Most participants were female (68.2%) and urban residents (85.7%). With respect to education, 88.6% had completed higher education (i.e., university), while 7.8% had only completed high school. The majority of participants were university students (81.7%), followed by smaller proportions of those who were employed (8.9%) or unemployed (2.8%). Based on self-reported engagement levels in mental health activities, 553 (46.7%) of participants were classified as being “engaged” versus 630 (53.2%) as “non-engaged” (Table 1). Across both groups, most participants were female (67.3% vs. 32.7%) residing in an urban area (84.4% vs. 15.6%) and single (2.3% vs 97.7%).

**Table 1:** Socio-demographic characteristics of study participants [number (percentage) or Mean ± standard deviation].

| Variable | Distribution N (%) |  |  |
| --- | --- | --- | --- |
|  | Non-Engaged<br>(N=630) | Engaged<br>(N=553) | Overall<br>(N=1183) |
| **Age (Mean ± SD)** | 20.6 (1.96) | 20.4 (2.04) | 20.5 (2.00) |
| **Sex** |  |  |  |
| Female | 435 (69.0) | 372 (67.3) | 807 (68.2) |
| Male | 195 (31.0) | 181 (32.7) | 376 (31.8) |
| **Place of residence** |  |  |  |
| Rural | 83 (13.2) | 86 (15.6) | 169 (14.3) |
| Urban | 547 (86.8) | 467 (84.4) | 1014 (85.7) |
| **Marital status** |  |  |  |
| Married | 14 (2.2) | 13 (2.4) | 27 (2.3) |
| Single | 616 (97.8) | 540 (97.6) | 1156 (97.7) |
| **Education** |  |  |  |
| Higher education | 585 (92.9) | 463 (83.7) | 1048 (88.6) |
| Other | 45 (7.1) | 90 (16.3) | 135 (11.4) |
| **Professional status** |  |  |  |
| Employed | 44 (7.0) | 61 (11.0) | 105 (8.9) |
| Unemployed | 586 (93.0) | 492 (89.0) | 1078 (91.1) |
| **Well-being drivers (Mean ± SD)** |  |  |  |
| Learning competence | 25.8 (5.9) | 26.4 (6.4) | 26.1 (6.1) |
| Good health and optimum nutrition | 27.0 (6.2) | 28.1 (6.6) | 27.5 (6.4) |
| Agency resilience | 20.4 (4.2) | 20.6 (4.5) | 20.5 (4.3) |
| Connectedness, positive values and contribution to society | 25.6 (6.9) | 27.8 (6.8) | 26.6 (6.9) |
| Safety and supportive environment | 23.2 (5.4) | 23.3 (5.5) | 23.3 (5.4) |
| Awareness about mental health | 5.8 (1.43) | 5.8 (1.6) | 5.8 (1.5) |
| Attitudes towards mental health | 26.8 (9.2) | 31.2 (11.0) | 28.9 (10.3) |

Regarding the drivers of well-being, overall, youth considered “good health and optimum nutrition” and “connectedness, positive values, and contribution to society” as the most important domains for their well-being (27.5 ± 6.41 and 26.6 ± 6.96, respectively). In contrast, “agency and resilience” was perceived as less important to well-being, receiving the lowest score overall (20.5 ± 4.27) and across both groups. In addition, youth who were not engaged in mental health initiatives assigned less significance to “connectedness, positive values and contribution to society” and “learning competencies” compared to their engaged counterparts (25.6 ± 6.92 vs. 27.8 ± 6.82) and (25.8 ± 5.87 vs. 26.4 ± 6.41), respectively.

Overall, the mean score for “awareness about mental health” was similar across both groups (5.84 ± 1.51), whereas higher levels of interest in mental health were recorded in the engaged vs non-engaged group, 7.60 ± 2.62 and 8.41 ± 2.16 respectively. For attitudes towards mental health, more positive attitudes were found in the engaged group compared to the non-engaged group (31.2 ± 11.0 vs 26.8 ± 9.21) (Table 1).

### 3.2. Description and comparaison of clusters

Three distinct clusters emerged from the analysis, differing significantly in mean engagement scores (p<0.001) (Table 2). Cluster 3 (n=332) had the highest engagement scores (10.9 ± 6.25), compared with cluster 1 and 2, and was characterized by a greater proportion of males (53.3%) and urban residents (84.6%). This cluster also had the highest employment rate (12.0%) and a relatively high percentage of participants pursuing higher education (79.8%). Cluster 3 consistently rated the importance of the five well -being dimensions significantly higher than the other clusters, particularly in ‘“Connectedness and positive values and contribution to society” to positive values, where they scored the highest (30.0 ± 5.29). A striking difference was observed in attitudes towards mental health, with Cluster 3 scoring substantially higher (41.1 ± 6.47) compared to Cluster 1 (25.1 ± 7.58) and Cluster 2 (27.7 ± 9.60) (p<0.001). Cluster 2 had the lowest engagement scores (5.23 ± 5.35) and the lowest scores across all well-being domains. This cluster had the highest proportion of males (66.9%) and rural residents (28.0%) and included the lowest percentage of individuals pursuing higher education (75.4%). Cluster 1, while also showing relatively low engagement (5.39 ± 4.94), differed from Cluster 2 by having a higher proportion of females (83.6%) and urban residents (88.4%). This cluster scored moderately across well-being dimensions, falling between Cluster 3 and Cluster 2 in most measures (Table 2).

**Table 2.**
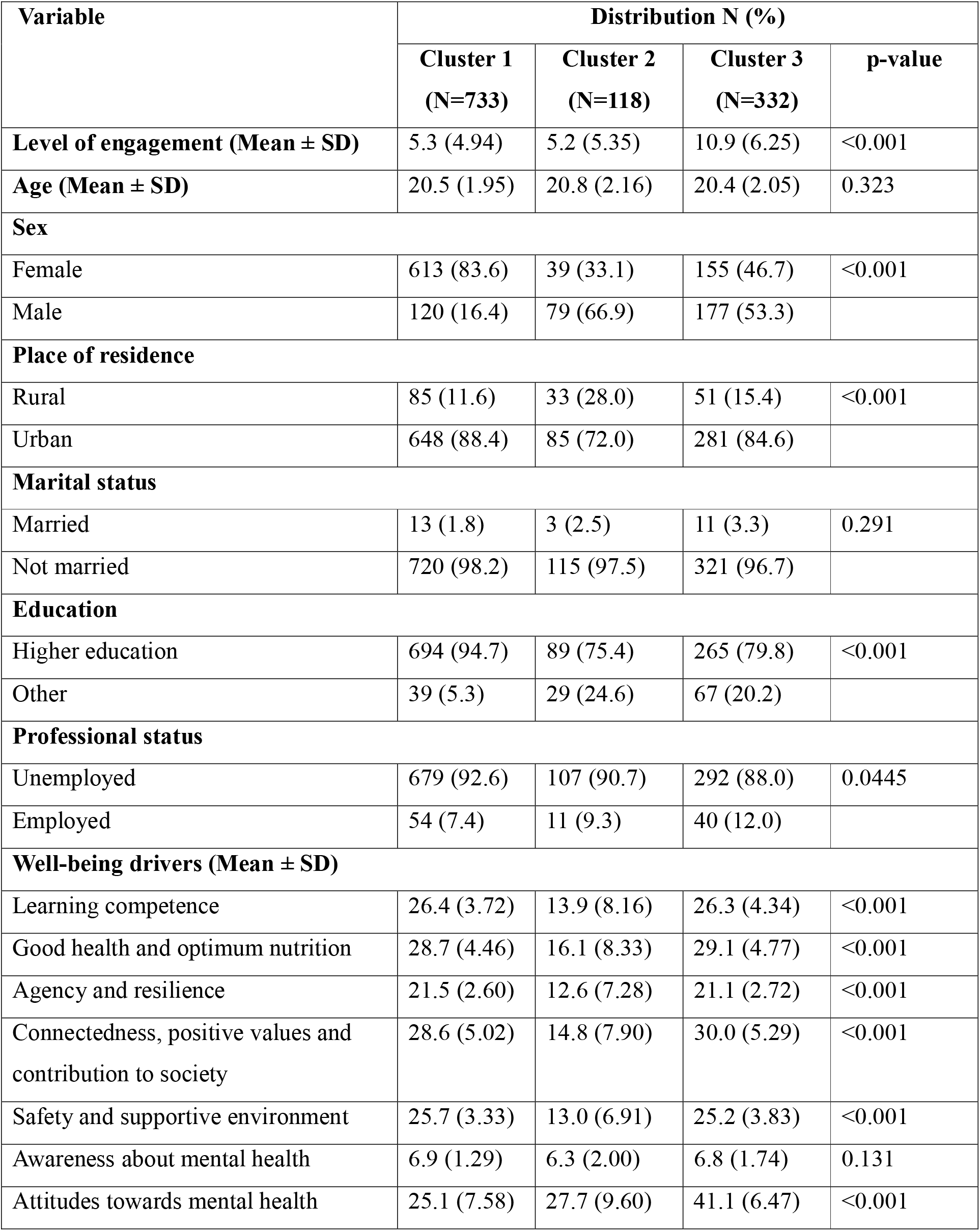
Profiles of youth by engagement clusters [number (percentage) or mean ± standard deviation].

### 3.3. Predictor factors of youth engagement in mental health

After multivariate analysis, youth who were more likely to be engaged included those with higher levels of education (OR = 2.23, 95% CI: 1.46-3.43, p<0.001), unemployed individuals (OR = 1.65, 95% CI: 1.04-2.64, p=0.035), those who rated “connectedness, positive values, and contribution to society” as highly important (OR = 1.07, 95% CI: 1.05-1.10, p<0.001), and those with more positive attitudes toward mental health (OR = 1.04, 95% CI: 1.03-1.05, p<0.001). In contrast, participants who placed greater importance on “safety and a supportive environment” were less likely to be engaged (OR = 0.94, 95% CI: 0.91-0.98, p=0.002) (Table 3).

**Table 3.** Predictor factors of youth engagement in mental health in Morocco

|  | Non-Engaged | Engaged | Crude OR (95% CI) | p-value | Adjusted OR<sup>a</sup> (95% CI) | p-value |
| --- | --- | --- | --- | --- | --- | --- |
| **Age** |  |  |  |  |  |  |
| Mean (SD) | 20.6 (2.0) | 20.4 (2.0) | 0.96 (0.91-1.02) | $p = 0.183$ | 0.97 (0.90-1.04) | $p = 0.373$ |
| **Sex** |  |  |  |  |  |  |
| Female | 435 (53.9) | 372 (46.1) | - |  | - |  |
| Male | 195 (51.9) | 181 (48.1) | 1.09 (0.85-1.39) | $p = 0.512$ | 0.90 (0.68-1.20) | $p = 0.484$ |
| **Place of residence** |  |  |  |  |  |  |
| Rural | 83 (49.1) | 86 (50.9) | - |  | - |  |
| Urban | 547 (53.9) | 467 (46.1) | 0.82 (0.59-1.14) | $p = 0.244$ | 0.81 (0.57-1.15) | $p = 0.236$ |
| **Marital status** |  |  |  |  |  |  |
| Married | 14 (51.9) | 13 (48.1) | - |  | - |  |
| Single | 616 (53.3) | 540 (46.7) | 0.94 (0.44-2.05) | $p = 0.883$ | 1.05 (0.45-2.47) | $p = 0.908$ |
| **Education** |  |  |  |  |  |  |
| Higher education | 585 (55.8) | 463 (44.2) | - |  | - |  |
| Other | 45 (33.3) | 90 (66.7) | 2.53 (1.74-3.71) | $p < 0.001$ | 2.23 (1.46-3.43) | $p < 0.001$ |
| **Professional status** |  |  |  |  |  |  |
| Unemployed | 586 (54.4) | 492 (45.6) | - |  | - |  |
| Employed | 44 (41.9) | 61 (58.1) | 1.65 (1.10-2.49) | $p = 0.015$ | 1.65 (1.04-2.64) | $p = 0.035$ |
| **Learning competence** |  |  |  |  |  |  |
| Mean (SD) | 25.8 (5.9) | 26.4 (6.4) | 1.01 (1.00-1.03) | $p = 0.134$ | 1.00 (0.97-1.04) | $p = 0.864$ |
| **Good health and optimum nutrition** |  |  |  |  |  |  |
| Mean (SD) | 27.0 (6.2) | 28.1 (6.6) | 1.03 (1.01-1.05) | $p = 0.005$ | 1.02 (1.00-1.05) | $p = 0.087$ |
| **Agency and resilience** |  |  |  |  |  |  |
| Mean (SD) | 20.4 (4.3) | 20.6 (4.5) | 1.01 (0.99-1.04) | $p = 0.398$ | 0.97 (0.93-1.02) | $p = 0.304$ |
| **Connectedness and positive values** |  |  |  |  |  |  |
| Mean (SD) | 25.6 (6.9) | 27.8 (6.8) | 1.05 (1.03-1.07) | $p < 0.001$ | 1.07 (1.05-1.10) | $p < 0.001$ |
| **Safety and supportive environment** |  |  |  |  |  |  |
| Mean (SD) | 23.2 (5.4) | 23.3 (5.5) | 1.00 (0.98-1.03) | $p = 0.641$ | 0.94 (0.91-0.98) | $p = 0.002$ |
| **Awareness about mental health** |  |  |  |  |  |  |
| Mean (SD) | 5.8 (1.4) | 5.8 (1.6) | 1.00 (0.93-1.08) | $p = 0.903$ | 0.99 (0.91-1.07) | $p = 0.749$ |
| **Attitudes towards mental health** |  |  |  |  |  |  |
| Mean (SD) | 26.8 (9.2) | 31.2 (11.0) | 1.04 (1.03-1.06) | p<0.001 | 1.04 (1.03-1.05) | p<0.001 |

<sup>a</sup>Odds ratios adjusted for for age, gender, residency, marital status, education, occupation, score of good health and optimum nutrition, score of connectedness, positive values, and contribution to society, score of safety and a supportive environment, score of learning, competence, education, skills, and employability, score of agency and resilience, score of awareness about mental health, and score of attitudes towards mental health.

### 3.4. Support and resources needed to engage in mental health initiatives

When asked about the resources needed to engage in mental health initiatives, the most commonly identified form of support was access to mental health professionals or counsellors (63.9%) (Figure 1). In addition, 42.4% highlighted the need for information and education about mental health issues. While not the primary concern for most, a significant proportion of participants also pointed to the importance of a safe space to discuss mental health issues (36.6%), support from friends and family (36.8%), financial assistance for mental health services (30.2%), and policy changes to improve access to mental health services (24.7%).

**Figure 1.**
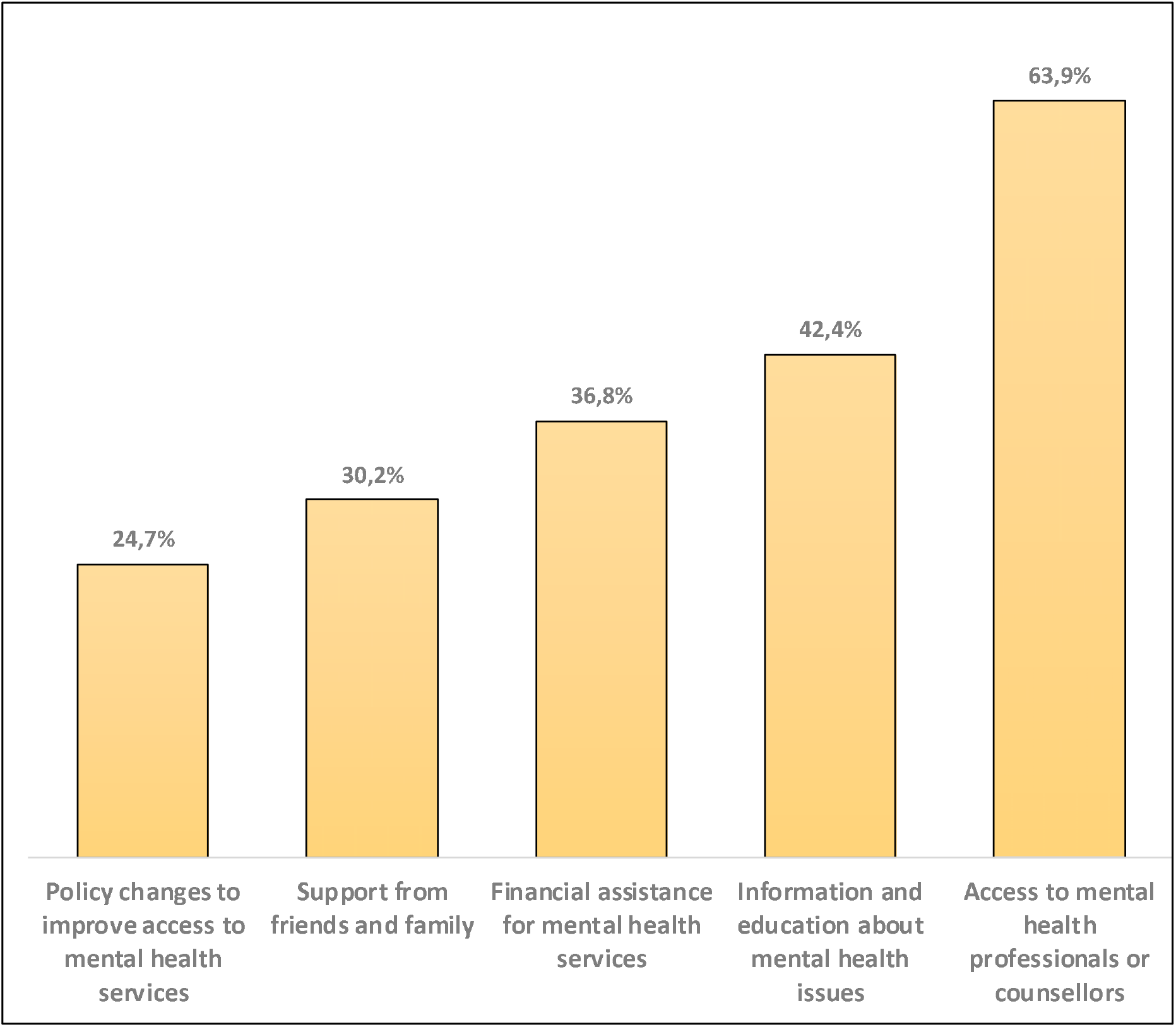
Support and resources needed to promote youth engagement in mental health (N=1,183)

## Discussion

This nationwide cross-sectional study among Moroccan youth aged 18 to 24 explored the predictors of youth engagement in mental health as well as the resources and support needed to promote it. The findings reveal that youth who are highly engaged in mental health initiatives typically exhibit strong positive attitudes toward mental health, value connectedness, and are more likely to be unemployed and pursuing higher education. Regarding support needs, many participants indicated that accessing mental health professionals or counselors is essential. The study highlights that mental health engagement is influenced by a combination of individual factors, such as education, employment status, and personal attitudes, and also perceived importance of contextual factors like environmental safety, social connectedness, positive values.

The study findings showed that higher education is a key driver of youth engagement in Morocco, reflecting broader evidence that education fosters the cognitive, social, and economic competencies necessary for meaningful participation. In Morocco, structural barriers disproportionately affect youth from low socio-economic backgrounds, young women and youth from rural communities, severely limiting their opportunities to pursue higher education (31). Given UNICEF’s emphasis on the pivotal role of schools in shaping adolescents’ life skills (32), and the country’s NEET (Not in Employment, Education, or Training) rate hovering at 26%—with young women being twice as likely to be in this category (34)—expanding access to quality education should be a central priority for policymakers. At the same time, our findings indicate that unemployed youth tended to be more engaged than their employed peers. This may reflect that unemployed youth, may have more unoccupied time, or actively seek mental health resources to better cope with the stresses of job hunting. Hence, targeted mental health programs and community initiatives tailored to unemployed youth could channel this heightened engagement into skill-building and resilience. Overall, these results highlight the need for multifaceted policies that simultaneously expand educational opportunities and as well as seize opportunity of unoccupied time during youth transitional phases to foster youth engagement in mental health activities in Morocco.

Another key finding in this study is that participants who perceived “safety and supportive environment” of high importance tended to be less engaged. This paradoxical insight could stem from two factors. First, youth that value a “safer and supportive environment” may struggle to find it in their daily lives limiting their ability to be more engaged. Alternatively, youth who may experience safer environments may not feel compelled to participate in mental health initiatives or may not have enough incentives to step outside of their comfort zones. The 2016 Moroccan GSH Survey conducted revealed that 16% of students had considered suicide in the year prior, with higher rates among girls (17.9% girls vs. 14% boys) (10) and 13.5% had used tobacco products before the age of 14 (10). These statistics reinforce the urgent need to create supportive environments that can foster healthier developmental outcomes. Globally, initiatives such as the “Supporting the Whole Student” offer effective strategies for creating more supportive educational environments. Therefore, adapting such approaches in Morocco could reduce exposure to violence and discrimination, thereby strengthening youth engagement and promoting better mental health outcomes (20).

“Connectedness, positive values and contribution to society” was rated of high importance by engaged youth in mental health and was statically significant in our study. Social connectednesss is essential for psychological well-being, with research linking it to reduced mortality risk and better mental health outcomes. A 2024 meta-analysis showcased that school connectedness significantly improved adolescent well-being, highlighting that adolescent with supportive peers and teachers show reduced mental health risks and higher resilience (21) even during the COVID-19 pandemic (22). Additionally, a large international study of 33,269 youth across 25 countries (International Survey of Children’s Well-Being, 2016–2019) underscored the role of positive teacher connections in both academic and psychosocial outcomes (23) while other studies emphasized the importance of investing in programs that strengthen connectedness with parents, peers, schools, and community (24). Moroccan youth, especially engaged youth in mental health echoed the importance of “Connectedness, Positive values and contribution to society” to promote engagement in youth.

Another important finding of this study is that participants with more positive attitudes towards mental health tended to be more engaged, joining the results in the global literature highlighting the link between mental health literacy and active involvement (25). Nearly half the participants requested additional resources to improve their mental health knowledge in this study, suggesting that limited literacy may hinder engagement. This emphasizes the need for youth-driven, culturally sensitive interventions that dismantle misconceptions, foster favorable attitudes, and empower youth to take part in mental health initiatives (26). Moreover, such programs must focus on directly addressing stigma to ensure adolescents develop the self-efficacy needed for sustained engagement. Considering the WHO’s (27) emphasis on early interventions to promote mental well-being, future endeavors should tailor educational and extracurricular activities to enhance mental health awareness (28). By doing so, stakeholders can expect a robust increase in both the quality and longevity of youth participation in mental health–related efforts.

The strong demand for mental health professionals among participants underscores the universal need for accessible counselling services. Countries worldwide—regardless of income level—are still in the process of developing their mental health systems, facing deep-rooted structural and social barriers (29). Meanwhile, the growing popularity of school-based interventions may offer a promising avenue for reaching young people more effectively, integrating mental health support into everyday environments (26,30). By reducing stigma, addressing logistical barriers, and cultivating a culture of help-seeking, such programs can ensure students receive timely and sustained mental health support. Given the responses from participants, it is imperative to invest in youth mental health services within the Moroccan context to effectively address the needs of young people.

To the best of our knowledge, this is the first study in Morocco to explore youth engagement in mental health including a large sample of participants across the 12 regions of Morocco. However, this study has some limitations. First, the cross-sectional design prevents drawing causal conclusions due the limited sample size. Although a stratified random sampling was used, our sample did represent mostly students vs non schooled, mostly from urban areas versus rural, and mostly unemployed vs employed participants which may not capture the full diversity of the target population. Hence, this study design reduces our ability to generalise these findings. Second, voluntary participation may have introduced selection as more interested individuals with specific interests or characteristics could have been overrepresented. Third, reliance on self-reported questionnaires raises the possibility of reporting bias and inaccuracies. We believe that future research should replicate these methods in larger, more diverse, and randomized samples to enhance validity and applicability.

## Conclusions

Promoting youth engagement in mental health activities in Morocco requires integrated policies that prioritize social connectedness, mental health literacy and access to mental health support. Community-based and school initiatives should aim at strengthening relationships among youth, peers, families, and educators to foster a sense of belonging. Expanding access to mental health professionals particularly in under-resourced areas, could address both underlying mental health illnesses and boost well-being and engagement. Together, these measures can empower Moroccan youth to become robust contributors to both mental health initiatives and their broader communities.

## Data Availability

The datasets used and/or analysed during the current study available from the corresponding author on reasonable request.

## Declarations

### Ethics approval

Approval for the study protocol was granted by the Ethics Committee of the Faculty of Medicine and Pharmacy of Tangier under number (AC470C/2023).

### Consent for publication

Obtained from all participants

### Competing interest

None to declare

### Funding

This study was financially supported by Grand Challenges Canada.

### Authors’ contribution

Conceptualization, W.F, B.O, and M.K, methodology, W.F. and B.O validation, B.I and M.K, formal analysis, O.B and B.I. data curation, W.F and B.O writing—original draft preparation, W.F, and B.O.; writing—review and editing, W.F, B.O.,, M.K and Z.A, and B.O supervision, M.K.; All authors have read and agreed to the published version of the manuscript.

## Acknowledgments

The authors would like to thank the young people who participated in this study as well as the Being Initiative for their support and funding to implement this nationwide research project. The Being Initiative is a youth mental health fund hosted by Grand Challenges Canada, with partnership of Fondation Botnar, The UK’s National Institute for Health and Care Research (NIHR), Orygen, the Science for Africa Foundation, and United for Global Mental Health.

## Notes

### Competing Interest Statement

The authors have declared no competing interest.

### Author Declarations

the Ethics Committee of the Faculty of Medicine and Pharmacy of Tangier gave ethical approval for this workunder number (AC470C/2023).

