## Supplemental file for "Drivers of youth engagement in mental health in Morocco: findings from a nationwide cross-sectional survey"

**Mental Health and Well-being Drivers, Engagement and Willingness to Take Action among Moroccan Youth**

***Survey***

**I. Socio-demographic and general information**

**How old are you?**

- 18
- 19
- 20
- 21
- 22
- 23
- 24

**Gender**

- Female
- Male

**Place of residence**

- Urban
- Rural

**City of residence:** ……………………..

**Phone number:** …………….…

**What is your marital status?**

- Single
- Married
- Widow
- Divorced or separated

**What is the highest level of education you have attained?**

- Not enrolled in school
- Primary
- Secondary
- High school
- Higher education/ Professional training

**Your current occupation:**

- Student
- Employed
- Unemployed
- Not in Education, Employment, or Training

**II. Mental health and well-being drivers**

**How important are the following elements to your well-being ?**

**1. Good health and optimum nutrition**

|  | **Extremely important** | **Very important** | **Moderately important** | **Slightly important** | **Not important at all** |
| --- | --- | --- | --- | --- | --- |
| Having a varied and balanced diet |  |  |  |  |  |
| Sleeping well at night |  |  |  |  |  |
| Practicing in sports and exercise |  |  |  |  |  |
| Being satisfied with the way I look |  |  |  |  |  |
| Being physically healthy |  |  |  |  |  |
| Having access to information about health |  |  |  |  |  |
| Having access to quality health services |  |  |  |  |  |
| Having access to green spaces |  |  |  |  |  |
| Breathing clean air |  |  |  |  |  |

**2. Connectedness, positive values, and contribution to society**

|  | **Extremely important** | **Very important** | **Moderately important** | **Slightly important** | **Not important at all** |
| --- | --- | --- | --- | --- | --- |
| Having a good relationship with my parents and family |  |  |  |  |  |
| Having a positive relationship with my teachers/professors/employer |  |  |  |  |  |
| Having a positive relationship with my peers/ colleagues |  |  |  |  |  |
| Feeling a sense of belonging in my community, school or work environment |  |  |  |  |  |
| Having an adult person in my life who I can trust |  |  |  |  |  |
| Feeling accepted, respected and valued by others |  |  |  |  |  |
| Participating in social and cultural activities in the community or at school /university or work |  |  |  |  |  |
| Having the chance to participate in decision-making and having my ideas valued and respected. |  |  |  |  |  |
| Getting the chance to learn about empathy, make friends, and become more understanding |  |  |  |  |  |

**3. Safety and a supportive environment**

|  | **Extremely important** | **Very important** | **Moderately important** | **Slightly important** | **Not important at all** |
| --- | --- | --- | --- | --- | --- |
| Not being exposed to violence *(including bullying, online harassment, physical, sexual, verbal abuse and emotional violence)* |  |  |  |  |  |
| Feeling safe in my daily life, whether at home, in my neighborhood, online, or at school or work. |  |  |  |  |  |
| Being treated like others and without discrimination |  |  |  |  |  |
| Feeling safe to express myself and be who I am |  |  |  |  |  |
| Having essential needs such as food, water, a place to live, warmth, clothing, and feeling safe and secure |  |  |  |  |  |
| My personal information is protected and is not shared without my permission |  |  |  |  |  |
| Having access to leisure activities and personal development opportunities |  |  |  |  |  |

**4. Learning, competence, education, skills, and employability**

|  | **Extremely important** | **Very important** | **Moderately important** | **Slightly important** | **Not important at all** |
| --- | --- | --- | --- | --- | --- |
| Getting to go to school and having chances to keep learning, whether in a classroom or through other kinds of learning |  |  |  |  |  |
| Getting help to stay motivated and keep learning, |  |  |  |  |  |
| Having chances to build the tools and skills to succeed |  |  |  |  |  |
| Having self-confidence and feeling that I can do things well |  |  |  |  |  |
| Learning practical skills for work |  |  |  |  |  |
| Working in jobs and businesses appropriate for my age |  |  |  |  |  |
| Being satisfied with my learning and skills |  |  |  |  |  |
| Believing in myself and my ability to reach my learning goals |  |  |  |  |  |

**5. Agency and resilience**

|  | **Extremely important** | **Very important** | **Moderately important** | **Slightly important** | **Not important at all** |
| --- | --- | --- | --- | --- | --- |
| Feeling independent and capable of making my own decisions |  |  |  |  |  |
| Feeling empowered to accomplish things and having confidence in myself (whether with friends, family, or in decision-making |  |  |  |  |  |
| Having hope and optimism towards the future |  |  |  |  |  |
| Having a sense of purpose in my life |  |  |  |  |  |
| Having opportunities to develop the ability to handle the challenges in life both now and in the future |  |  |  |  |  |
| Having chances to reach my full potential now and later in life. |  |  |  |  |  |

**III. Awareness about mental health:**

|  | **True** | **False** | **Don’t know** |
| --- | --- | --- | --- |
| Mental health is a component of health or like any other diseases |  |  |  |
| Suicidal ideation or suicidal attempt is one of the psychological problems |  |  |  |
| Psychological problems or mental illness can start at a very early age |  |  |  |
| Because of bullying or abuse one can develop psychological problems and mental disorders |  |  |  |
| Middle-aged individuals are unlikely to develop mental disorders |  |  |  |
| Patients with mental disorders are not always sad |  |  |  |
| A person with depression feels very miserable |  |  |  |
| Doing something enjoyable helps to improve mental health |  |  |  |

**IV. Attitudes about mental health:**

|  | **Completely agree** | **Agree** | **Undecided** | **Disagree** | **Strongly disagree** |
| --- | --- | --- | --- | --- | --- |
| I might be judged by others if I talk about my mental health. |  |  |  |  |  |
| I don't always know where to find help when I need it. |  |  |  |  |  |
| I believe there aren’t enough investment or resources in mental health programs. |  |  |  |  |  |
| I sometimes can't get to places where mental health support is available because it's too far or costs too much. |  |  |  |  |  |
| I might have trouble finding help if people around me speak different languages or have different backgrounds. |  |  |  |  |  |
| I don’t understand what mental health really means or why it's important. |  |  |  |  |  |
| I worry that if I ask for help or try to help others, I might be treated unfairly or judged by others. |  |  |  |  |  |
| I have a busy schedule with school/university/workplace, so I might not have much time to get involved in mental health activities. |  |  |  |  |  |
| I might feel like I'm the only one who cares about mental health, which can make me feel alone. |  |  |  |  |  |
| I might have had a bad experience with mental health services before, which makes me hesitant to try again. |  |  |  |  |  |
| What my parents or family think can affect whether I get involved in mental health activities. |  |  |  |  |  |
| Mental health-related activities are not organized in my environment (school, university, neighborhood, workplace,...). |  |  |  |  |  |
| I might not have role models or people to look up to when it comes to getting involved in mental health. |  |  |  |  |  |

**V. Youth engagement**

**To which extent to do you agree with the following:**

|  | **Always** | **Very often** | **Sometimes** | **Rarely** | **Never** |
| --- | --- | --- | --- | --- | --- |
| How often do you engage in activities that promote mental health and well-being? |  |  |  |  |  |
| I am involved in the design of programs and activities related to mental health |  |  |  |  |  |
| I am involved in the implementation of mental health programs. |  |  |  |  |  |
| I am involved in keeping track of and evaluating mental health programs |  |  |  |  |  |
| I am actively involved in raising awareness about mental health |  |  |  |  |  |
| I participate in groups that make decisions about mental health. |  |  |  |  |  |

**What support would you need to be more engaged in initiatives or activities related to mental health for young people in Morocco?**

- Access to mental health professionals or counsellors
- Support from friends and family
- Information and education about mental health issues
- A safe space to talk about mental health issues
- Financial assistance for mental health services
- Policy changes to improve access to mental health services
- Other (please specify)
